# Addressing unmet social needs using a health navigator for patients at a major metropolitan hospital in Australia: a mixed-methods feasibility study

**DOI:** 10.1101/2024.04.02.24305238

**Authors:** K Neadley, C Shoubridge, A Smith, S Martin, M Boyd, C Hocking

**Affiliations:** University of Adelaide, South Australia, SA 5000; Northern Adelaide Local Health Network, South Australia; Australian Institute of Family Studies, Southbank, VIC 3006

## Abstract

**Introduction:** Integrating health and social care to address unmet social needs is an emerging priority for health systems worldwide. Screening and referral interventions for unmet social needs in healthcare settings have shown promising results. Most screening and referral interventions are implemented in primary care, despite evidence that disadvantaged populations face substantial barriers to accessing such care. There are few social care interventions in hospital settings. To address this gap, we designed a hospital-based intervention screening an outpatient population for unmet social needs and using a Health Navigator to provide referrals and follow-up to appropriate community and government resources. Here we present a protocol for a feasibility and acceptability study of a hospital-based Health Navigator intervention.

**Methods and Analysis:** We will conduct a single-centre study to explore the feasibility and acceptability of screening and referral for unmet social needs for patients attending an outpatient cancer clinic at a major metropolitan hospital serving a disadvantaged population in South Australia. Eligible participants are 18 years of age or older receiving treatment at the Northern Adelaide Cancer Centre, with an expected prognosis of minimum six months. Eligible participants will be asked to complete unmet social needs screening and baseline assessments. Participants with unmet social needs who request assistance will be connected with a Health Navigator (HN). The HN will work with participants to prioritise their needs and provide referrals to community and government services with follow-up of over six months from enrolment. Post-HN intervention, all participants will be asked to complete repeat unmet social needs screening and repeat assessments. The primary criteria for determining feasibility success are: 1) recruitment rates will be successful if 80% of eligible participants agree to unmet needs screening, 2) intervention uptake will be successful if 80% complete follow-up, 3) reasons for not completing intervention and 4) participant and clinician acceptability of the intervention. Secondary outcomes include changes to clinical measures such as coping capacity, quality of life and patient-reported experience measures. Thematic analysis will be applied to focus groups with clinicians and participants to assess intervention acceptability. Secondary clinical outcomes will be reported as effect size estimates for future trial. As feasibility studies are designed to test whether an intervention is appropriate for larger studies, rather than finding specific associations or outcomes, no sample size calculation is necessary. Study findings will be used to optimise recruitment and intervention components, and develop suitable outcome measures for larger, randomized studies.

**Ethics and Dissemination:** The protocol has ethical approval from the Central Adelaide Local Health Network Human Research Ethics Committee (approval ID: 16448).

Trial registration: ACTRN12622000802707p

Protocol date and version: 07 June 2022, V1

**Strengths and limitations of this study:**

- Most screening and referral interventions for unmet social needs occur in primary care, despite disadvantaged populations facing significant barriers to accessing primary care. This study takes place in a hospital setting.
- There are few interventions for unmet social needs in Australia. This study is a valuable contribution to screening and referral research in Australia.
- This study employs a screening tool for unmet needs co-designed with clinicians and community, and is one of few validated screening tools for unmet needs.
- The study population is limited to people living with cancer who experience substantial healthcare needs and treatment adverse effects. Findings are unlikely to be representative of the general population.

## Introduction

### Social determinants of health: upstream to the immediate

Health and wellbeing is inextricably linked to social circumstances. Social determinants of health (SDoH; e.g. housing, neighbourhood & physical environment, safety, food availability, social connection) are drivers of health and wellbeing[1]. SDoH are reinforced at the policy level, downstream of which are the social risks affecting individuals: housing and financial stress, food insecurity, social isolation and interpersonal violence[1, 2]. Over forty years ago the Whitehall studies reported the impact of these social risks: a strong inverse correlation between socioeconomic status and health outcomes[3, 4]. This ‘social gradient’ in health persists today, across all economies, countries and healthcare systems[1]. This inequity in health was particularly defined during the COVID-19 pandemic[5]. COVID-19, aging populations and the shift in global burden of disease to chronic illness, has created momentum for many countries to include action on unmet social needs as a health policy priority[6].

### Health and social care integration: addressing the ‘causes of the causes’

An increased understanding of the social context of health and transitions to value-based payment systems has increased social care integration in healthcare systems[6]. One mechanism of health and social care integration has been rapidly implemented, particularly in the US and UK: screening and referral interventions for unmet social needs[7, 8]. These interventions comprise three steps: 1) Screening for unmet social needs (e.g. housing stability, financial stress etc) in a healthcare setting using a dedicated screening tool for unmet social needs, 2) linking patients reporting unmet needs to appropriate government and community resources and 3) patient follow-up. Some interventions employ a Health Navigator (HN-also known as a ‘link worker’ or ‘community healthcare worker’) to assist patients with connections to resources, and provide advocacy and psychosocial support throughout follow-up in the community[8].

Currently, evidence for these interventions is mixed due to heterogeneity in study design and reported outcomes[7, 9]. HN training and role descriptions vary, as do the duration and frequency of follow-up in different settings[8]. Some studies report improvements in health outcomes[10] and resolution of unmet social needs[11] with an HN, while others report no change in access to resources[12], unmet needs or health outcomes between groups with an HN and controls[13]. Although these interventions are designed to assist the most marginalised and medically-underserved populations, reported rates of uptake of screening and referral interventions are lower in low-resource healthcare settings compared to uptake in more advantaged settings[14]. Most HN interventions take place in primary care[6], despite evidence that disadvantaged populations face substantial barriers to accessing this care[15] and are more likely to present to hospital[16]. In fee-for-service primary care systems such as Australia, care that can be reimbursed is prioritised, leaving clinicians without financial incentive to address unmet social needs. Furthermore, the acceptability of HN interventions is debated. Evidence suggests between 40-60% of participants are likely to respond to unmet needs screening, and that participants with higher reported unmet needs are less likely to request assistance with identified needs[14]. This may be due to patients’ fears of mandatory reporting requirements, or healthcare providers’ reported discomfort asking sensitive social questions and lack of knowledge of community resources to provide appropriate referrals[14].

Populations living with cancer are particularly affected by employment and financial instability as treatment requires protacted contact with the healthcare system, and adverse treatment effects can reduce earning capacity[17]. This change in financial circumstances directly related to cancer treatment and care is referred to as ‘financial toxicity’[18]. In Australia, one in four cancer patients pay more than AUD 10,000 out-of-pocket costs every two years[19]. As treatment and care can extend for multiple years, the impact of financial toxicity can be enduring, with ongoing implications for patients’ carers and dependents[20].

To our knowledge, HN intervention research in Australia is limited. Here we present a protocol for a pilot examining the feasibility and acceptability of an HN intervention to address unmet social needs for populations accessing cancer care in an Australian hospital setting. We hypothesise that a Health Navigator intervention for unmet social needs in an outpatient cancer centre will be a feasible intervention and acceptable to participants and clinicians.

To assist with integration of screening for unmet needs in this study we co-designed the screening tool for unmet social needs with local community receiving cancer treatment and clinicians to maximise screening acceptability.

### Aims and Objectives

The primary aim of this study is to examine the feasibility and acceptability of an HN intervention in an outpatient hospital population living with cancer. We will assess feasibility through rates of recruitment, intervention uptake and intervention completion. Acceptability will be assessed through focus groups and brief surveys with clinicians and participants. Our secondary aim is to explore the impact of the HN intervention on participants’ experience of care, quality of life and ability to cope with cancer.The primary objective of the study is to evaluate and optimise the HN research process, including data collection and maintenance, and to refine intervention procedures and outcome measures for future studies.

## Methods and Analysis

### Setting

The study will be conducted in northern Adelaide, South Australia. Participants will be recruited from one hospital outpatient cancer care setting serving the target population. This site has been selected as it serves a highly disadvantaged population[21], and there is evidence that social risks such as financial instability, food and housing insecurity are highly prevalent[22]. Clinicians frequently report patients undergong chemotherapy living in their cars, or missing treatments to work unstable jobs.

The study is planned to commence 8 August 2022 and complete 29 February 2024.

### Participants and sample size

Inclusion criteria are:

- Able to provide written, informed consent in English
- Adults ≥18 years old with any cancer diagnosis
- Receiving care or follow-up at the Northern Adelaide Cancer Centre

Exclusion criteria are:

- Patients deemed by oncologist to have expected survival <6 months
- Patients deemed by the Principal Investigator to not be capable of complying with protocol procedures

As this is a feasibility study designed to inform appropriate recruitment strategies and outcome measures for future randomized trials, we did not calculate a specific sample size.

### Outcome measures

Our primary aim is to determine intervention feasibility, which will be evaluated using:

1. Recruitment rates, will be successful if 80% of eligible participants agree to unmet needs screening
2. Intervention uptake, will be successful if 80% % of participants with unmet social needs consent to take part in the HN service
3. Intervention completion, will be successful if 80% of participants with unmet social needs complete intervention
4. Reasons for non-participation and failure to complete the intervention
5. Brief questionnaires sent to all participants and clinicians to explore intervention feasibility and acceptability
6. Focus groups with participants and clinicians to explore acceptability of the HN intervention

Other feasibility studies exploring screening and referral for unmet social needs in non-cancer populations have used a threshold for success of 80-90%[23]. We have chosen to lower our threshold to 80% to accommodate the adverse treatment effects[24] and psychosocial complications populations living with cancer may experience[25].

Our secondary aim is to optimise outcome selection for a larger randomized study. All participants (with and without needs) will complete baseline unmet needs screening[26], Coping with Cancer (CBI-B)[27], Functional Assessment of Cancer Therapy (FACT-G)[28] and Australian Hospital Patient Experience Questionnaire Set (AHPEQS)[29] measures. After follow-up is completed these measures will be repeated. Pre/post analysis of these measures will be performed for participants with and without needs and changes in measures will be compared between groups.

### Study procedure

As this is an efficacy-focused feasibility study, researchers will be responsible for recruitment. Clinicians will refer eligible participants to the researcher who will obtain written, informed consent and conduct baseline clinical measures (FACT-G, CBI-C and AHPEQS), and unmet needs screening using the Unmet Needs Screening Tool (UNST)[26]. The researcher will collect participant contact and demographic details. We will ask eligible participants who decline participation reasons for their decision.

If the participant has ≥1 unmet need, they will be referred to the HN. The researcher will arrange a time for the participant to meet with the HN for an initial meeting. Participants without any identified unmet needs will not be referred to the HN for intervention. During the initial HN meeting, the HN will work collaboratively with the participant to prioritise their three most important unmet needs (if they have more than one). This will inform the case management plan and referral to community-based resources. If the participant requires any assistance with contacting the appropriate community resource, the HN will help by making the phone call for them, organising their meeting time, organising transport to get to the meeting and if requested, attend meetings with the participant. All these options, requests and case management plans will be recorded in the study database and will be provided to participants in hardcopy or via email for their own reference. All the information regarding community resources and appropriate contact details, including crisis support resources, will be given to the participant by the HN in an information pack. The HN will end the initial meeting once the participant has decided which service provider will be contacted first, the goal for meeting with the service, the timeframe for engaging with the service and their desired endpoint. Participants will be able to call the HN at any time during their work hours by study phone if they require assistance or clarification. The HN will arrange a time for follow-up at the participant’s convenience. Outside of working hours, there will be a message system that provides HN working hours and allows participants to leave their contact details for follow-up during working hours.

The first follow-up contact will be offered within 7-14 days of baseline assessment, and approximately every 4 weeks thereafter until study completion (24 weeks post-assessment) or participants decline further assistance. This follow-up schedule can be adjusted by the participant as preferred. At each follow-up contact point, the participant will notify the HN of progress towards resolving their unmet need(s). If their needs have been met, this will be classified as ‘resolved’, and the process will be repeated for other identified unmet needs. If more time is required with the same service this will be classified as ‘engaged’ and revisited in the next follow-up call. If the resource was not useful or unable to meet the participants’ needs, this will be classified as ‘failed’ and another resource will be identified by the HN to assist the participant. The HN will also ask participants why the referral was unsuccessful. Participants will be classified as ‘lost to follow-up’ if they can no longer be reached after three attempted contacts by the preferred mode of conduct. These classifications and call attempts will be captured during each follow-up call.

Post-HN intervention, participants and clinicians will be asked to take part in separate focus groups to explore their views of the HN intervention, and barriers and enablers to screening for unmet social needs in the outpatient hospital setting.

### Data Collection

Data will be collected and managed using REDCap electronic data capture tools hosted at the University of Adelaide. REDCap (Research Electronic Data Capture) is a secure, web-based software platform designed to support data capture for research studies, providing 1) an intuitive interface for validated data capture; 2) audit trails for tracking data manipulation and export procedures; 3) automated export procedures for seamless data downloads to common statistical packages; and 4) procedures for data integration and interoperability with external sources.

### Data Analysis

All quantitative data will be analysed using descriptive statistics in STATA MP17 software. Qualitative data will be analysed using reflexive thematic analysis and NVivo 15 software. Criteria for feasibility success, outcomes and analyses are summarised in Table 2.

**Table 2:** Summary of objectives, outcomes, criteria for success of feasibility, and methods of analysis

| Aim | Objective | Outcome | Criteria for feasibility success | Analysis |
| --- | --- | --- | --- | --- |
| Primary | Feasibility of intervention | Recruitment rate | ≥80 % of eligible participants target sample recruited | Descriptive, percentage (95% CI) |
|  |  | Intervention uptake | ≥80 % of eligible participants take up intervention | Descriptive, percentage (95% CI) |
|  |  | Intervention completion | ≥80 % of participants complete intervention | Descriptive, percentage (95% CI) |
|  |  | Feasibility and Acceptability Intervention Measures | ≥80 % clinicians find the intervention feasible and acceptable | Descriptive, percentage |
|  |  | Reasons for intervention withdrawal |  | Categorical, percentage |
| Secondary | Clinical outcomes | Change in satisfaction with care (AHPEQS) | Intervention group will show improvement | Estimates of effects (95% CI) |
|  |  | Change in Coping with Cancer score |  | Estimates of effects (95% CI) |
|  |  | Change in quality of life (FACT-G) |  | Estimates of effects (95% CI) |

### Patient and Public Involvement

Participants were first involved in our research in the formulation and validation of the Unmet Needs Screening Tool (referred to as the screening tool hereafter). The screening tool was co-designed and qualitatively validated with participants from previous studies and clinicians working with the target population[26].

A previous validation study in the target population informed our approach to screening with the screening tool (under review, trial registration:ACTRN12620001326987). The study determined that most participants preferred to complete the UNST individually, without assistance. In this study, we will offer participants the option to complete the screening tool alone, without involvement from a researcher.

In the future, we hope to use the experience of these participants to further refine the screening tool. We will use participants’ experiences with the HN to inform future study design, processes, follow-up and refine the HN role for future implementation.

## Data Availability

All data produced in the present study are available upon reasonable request to the authors

## Ethics and Dissemination

The HNs will ask for consent to share participants’ contact details with appropriate community services, and inform participants of their responsibility to report any circumstances which may endanger themselves or those around them. If there is are concerns for safety of the participants or those around them, the HN will notify the Principal Investigator who will follow appropriate procedures. We expect the HN role to be challenging and will ensure specific measures are in place to address any concerns that may arise. This includes prioritising time every week for the HN to debrief about case load, case severity and provide feedback regarding study processes. Study results will be disseminated to participants, clinicians and community service providers, as appropriate. All investigators will be listed as authors on all publications.

## Authors’ Contributions

MB, SM, KN, CH, AS, CS were responsible for study design. All authors wrote and reviewed the protocol manuscript.

## References

1. CSDH, Closing the gap in a generation: health equity through action on the social determinants of health. Final Report of the Commission on Social Determinants of Health. 2008: Geneva.

2. Andermann, A., Screening for social determinants of health in clinical care: moving from the margins to the mainstream. Public Health Rev, 2018. 39: p. 19.

3. Marmot, M., et al., Employment grade and coronary heart disease in British civil servants. J Epidemiol Community Health, 1978. 32: p. 244–249.

4. Marmot, M., et al., Health inequalities among British civil servants: the Whitehall II study. The Lancet, 1991. 337: p. 1387–1393.

5. COVID-19 and the social determinants of health and health equity: evidence brief. 2021, World Health Organisation: Geneva.

6. Morse, D.F., et al., Global developments in social prescribing. BMJ Glob Health, 2022. 7(5).

7. Yan, A.F., et al., Effectiveness of Social Needs Screening and Interventions in Clinical Settings on Utilization, Cost, and Clinical Outcomes: A Systematic Review. Health Equity, 2022. 6(1): p. 454–475.

8. Sandhu, S., et al., Intervention components of link worker social prescribing programmes: A scoping review. Health Soc Care Community, 2022. 30(6): p. e3761–e3774.

9. Ruiz Escobar, E., S. Pathak, and C.M. Blanchard, Screening and Referral Care Delivery Services and Unmet Health-Related Social Needs: A Systematic Review. Prev Chronic Dis, 2021. 18: p. E78.

10. Berkowitz, S.A., et al., Addressing Unmet Basic Resource Needs as Part of Chronic Cardiometabolic Disease Management. JAMA Intern Med, 2017. 177(2): p. 244–252.

11. Gottlieb, L.M., et al., Effects of Social Needs Screening and In-Person Service Navigation on Child Health: A Randomized Clinical Trial. JAMA Pediatr, 2016. 170(11): p. e162521.

12. Lopez, M.A., et al., Social Needs Screening in Hospitalized Pediatric Patients: A Randomized Controlled Trial. Hosp Pediatr, 2023. 13(2): p. 95–114.

13. Mercer, S.W., et al., Effectiveness of Community-Links Practitioners in Areas of High Socioeconomic Deprivation. Ann Fam Med, 2019. 17(6): p. 518–525.

14. Kreuter, M.W., et al., How Do Social Needs Cluster Among Low-Income Individuals? Popul Health Manag, 2021. 24(3): p. 322–332.

15. Corscadden, L., et al., Factors associated with multiple barriers to access to primary care: an international analysis. Int J Equity Health, 2018. 17(1): p. 28.

16. Unwin, M., et al., Socioeconomic disadvantage as a driver of non-urgent emergency department presentations: A retrospective data analysis. PLoS One, 2020. 15(4): p. e0231429.

17. Gordon, L.G., et al., Reduced employment and financial hardship among middle-aged individuals with colorectal cancer. Eur J Cancer Care (Engl), 2017. 26(5).

18. Desai, A. and B. Gyawali, Financial toxicity of cancer treatment: Moving the discussion from acknowledgement of the problem to identifying solutions. EClinicalMedicine, 2020. 20: p. 100269.

19. Bygrave, A., et al., Australian Experiences of Out-of-Pocket Costs and Financial Burden Following a Cancer Diagnosis: A Systematic Review. Int J Environ Res Public Health, 2021. 18(5).

20. Chan, R., et al., Distinct financial distress profiles in patients with breast cancer prior to and for 12 months following surgery. BMJ Support Palliat Care, 2022. 12(3): p. 347–354.

21. Socio-Economic Indexes for Area (SEIFA): Local Government Area, SA1 Distributions, ABS, Editor. 2021: Canberra.

22. Neadley, K.E., et al., Capturing the social determinants of health at the individual level: a pilot study. Public Health Res Pract, 2021. 31(2).

23. Wahi, G., et al., Screening and addressing social needs of children and families enrolled in a pediatric weight management program: a protocol for a pilot randomized controlled trial. Pilot Feasibility Stud, 2022. 8(1): p. 129.

24. De Ruysscher, D., et al., Radiotherapy toxicity. Nat Rev Dis Primers, 2019. 5(1): p. 13.

25. Niedzwiedz, C.L., et al., Depression and anxiety among people living with and beyond cancer: a growing clinical and research priority. BMC Cancer, 2019. 19(1): p. 943.

26. Poirier, B.F., et al., Development of Social Determinants of Health Screening Tool (SDoHST): qualitative validation with stakeholders and patients in South Australia. Curr Med Res Opin, 2023. 39(1): p. 131–140.

27. Heitzmann, C.A., et al., Assessing self-efficacy for coping with cancer: development and psychometric analysis of the brief version of the Cancer Behavior Inventory (CBI-B). Psychooncology, 2011. 20(3): p. 302–12.

28. Peipert, J.D. and D. Cella, Bifactor analysis confirmation of the factorial structure of the Functional Assessment of Cancer Therapy-General (FACT-G). Psychooncology, 2019. 28(5): p. 1149–1152.

29. ACSQHC, Summary of Development and Testing of the AHPEQS, A.C.o.S.a.Q.i.H. Care, Editor. 2017: Sydney.

